# Treating Unmet Needs in Psychiatry (TUNE-UP): A Targeted Service Increases Outpatient Initiations of Clozapine

**DOI:** 10.1101/2025.04.04.25325236

**Authors:** Zmarak Ahmad Khan, Ioana Varvari, Valentina Mancini, Chambrez Zita Zauchenberger, Shashwati Kantor, Jack B Fanshawe, Sharon Musiiwa, Alexandra Pledge, Benjamin Pearce, Saik de la Motte, Digby Quested, Daniel Maughan, Philip McGuire, Oliver Howes, Toby Pillinger, Robert McCutcheon

## Abstract

**Background:** Up to one-third of patients with schizophrenia do not benefit from standard antipsychotic treatment – termed treatment-resistant schizophrenia (TRS). Clozapine is the only licensed treatment in TRS and is associated with better outcomes. However, it is underused, as its initiation is often limited by the need for inpatient admission, which is costly and unattractive to patients. Community clozapine titration services may address this.

**Aims:** To describe a targeted outpatient clinic (TUNE-UP) for TRS management and assess its impact on clozapine initiation rates.

**Method:** We reviewed clozapine titrations for patients under four community mental health teams in the United Kingdom from September 2021 to January 2025, noting whether titration occurred in inpatient or outpatient settings. The TUNE-UP clozapine clinic operated for 12 months (September 2023 to September 2024). Initiation rates during the TUNE-UP period were compared with rates when the service was unavailable using Poisson regression. Clinical outcomes were assessed using scales such as the Positive and Negative Syndrome Scale (PANSS) for symptom severity and the Social and Occupational Functioning Assessment Scale (SOFAS) for functioning.

**Results:** 61 individuals were commenced on clozapine. During the TUNE-UP clinic’s operation, community initiation rates increased to 11.0 per year (up from 1.33 per year when the service was unavailable), while inpatient initiations were similar (11.0 per year vs. 11.67 per year). Increases in total initiations (p = 0.048) and community initiations (p = 0.0003) were statistically significant. Patients seen by TUNE-UP had improvements in PANSS (mean baseline 63.3 (SD 18.3); mean improvement 20.5 (SD 12.2; p = 0.009)), and SOFAS (mean baseline 48.3 (SD 7.5); mean improvement 8.8 (SD 7.1, p = 0.028)).

**Conclusion:** A specialist community service was associated with a significant increase in clozapine initiations alongside improved clinical outcomes. This approach offers a clinically and cost-effective strategy to enhance treatment for TRS.

## Introduction

Schizophrenia is a psychiatric disorder consisting of positive, negative, and cognitive symptoms, and is associated with significant functional impairment.(1) Up to one-in-three patients have treatment-resistant schizophrenia (TRS).(2) TRS is defined as an inadequate response to treatment of adequate dose, duration, and concordance with two different antipsychotics.(3,4)

Clozapine is the only antipsychotic treatment that is specifically licensed for use in TRS, and it is associated with better outcomes compared with other antipsychotics in this patient population.(3–6) Despite guidelines advising treatment with clozapine at the earliest opportunity in TRS, there can be significant delays to initiating clozapine in clinical practice, (7–9) and this is associated with worse outcomes.(10)

Delays in clozapine initiation can result, in part, from the fact that initiation is often only available in an inpatient setting. This is typically an unattractive option for patients and incurs significant financial cost.(11,12) Community titrations of clozapine have previously been shown to be a safe and feasible approach, and are associated with significant reductions in cost, service use and symptom severity.(11) The impact of a specialist community initiation service on clozapine initiation rates in the community, however, has not yet been investigated.

We sought to investigate if a community clozapine titration service is associated with an increase in the rate of community clozapine titrations. We hypothesised that there would be an increase in community clozapine titrations in the time that this service was available, compared to periods when it was not. We also hypothesised a concurrent decrease in inpatient clozapine titrations during the period this service was available. We assessed key clinical outcomes at baseline and discharge, in patients titrated through this service, as a secondary objective.

## Methods

We performed a retrospective analysis of clozapine titration locations over 40 months (September 2021-January 2025). This period consisted of two years as a baseline prior to the service’s commencement, followed by the one year this service was operational, followed by the four-month period after its cessation. During 12 of these months (Year 3) this specialist community service, the Treating Unmet Needs in Psychiatry (TUNE-UP) clinic, was in operation.

### Objectives

The primary objective was to compare the rates of community clozapine titration in the year the TUNE-UP service was running to the two-years beforehand, and the four-month period afterwards. Secondary objectives included a comparison of combined (i.e. community and inpatient) clozapine titration rates, and investigation of the effect of clozapine initiation on clinical measures.

### Service description

The TUNE-UP clinic launched in September 2023 as a satellite service to four Oxford Health NHS Foundation Trust (United Kingdom) community psychiatry teams (three adult mental health teams (AMHTs) and one early intervention service (OEIS)). These teams serve a population of approximately 165,000 people and have a combined caseload of 1455 patients.

Patients can be referred by these teams to TUNE-UP for assessment and management advice of refractory positive, negative, and cognitive symptoms or physical health concerns. A comprehensive assessment is performed (see box 1), and a management plan is developed in collaboration with the patient. Following approval by the referrer, the TUNE-UP service then implements the proposed plan, including, but not limited to, community initiation of clozapine (see box 2, based on existing community clozapine initiation approaches).(13)

##### Box 1: The TUNE-UP clinic service model

###### From referral, within 3 weeks

- **Review** patient electronic notes and obtain collateral information.
- **Explore** and help patients articulate and formulate their goals.
- **Conduct** a comprehensive assessment including standardised rating scales:
  – Positive and Negative Syndrome Scale (PANSS).(14)
  – Brief Negative Syndrome Scale (BNSS).(15)
  – Calgary Depression Scale (CDS).(16)
  – Screen for Cognitive Impairment in Psychiatry (SCIP).(17)
  – Subjective Scale to Investigate Cognition in Schizophrenia (SSTICS).(18)
  – Sheehan Disability Scale (SDS).(19)
  – Social and Occupational Functioning Assessment Scale (SOFAS).(20)
  – Sleep Condition Indicator (SCI).(21)
- **Investigate** physical health via physical examination and clinical investigations including ECG and blood tests (prolactin, full blood count (FBC), erythrocyte sedimentation rate (ESR), thyroid function tests (TFTs), liver function tests (LFTs), urea and electrolytes (U&Es), bone profile, lipids profile, HbA1c, c-reactive protein (CRP), Troponin-I (for clozapine titrations), and, if indicated, psychotropic levels).

###### From investigation stage to management formulation stage, 1 week

- **Identify** treatment options for a range of symptom domains: positive symptoms, negative symptoms, affective symptoms, sleep, cardiometabolic health.
- **Develop** treatment strategies via a **Co-Care Approach Model (involving AMHT, GP and carers) aligned with patient goals**, such as side-effect informed treatment optimization(22), side-effects management and adjunctive therapies including psychosocial interventions.

###### Implementation (3-6 months)

- **Implement and monitor changes**
- **Longitudinal assessment** with repetition of clinical scales at discharge
- **Discharge** to AMHT or GP with short-term and long-term recommendations that align with patient goals.

##### Box 2: Community clozapine initiation procedures

###### After a patient meets the treatment resistance criteria, the following steps are conducted

→ Community initiation eligibility screen as per Maudsley Guidelines.(3)
→ Baseline investigations: ECG, FBC, Troponin-I, CRP, LFTs, U&Es, glucose, lipids, HBA1c.

###### Considering patient preference, a tailored titration plan weighing mental state, current psychotropics and prior trials/responses to clozapine is designed. A standard plan is

→ Week 1: start at 6.25mg and double dose every other day reaching 50mg at the end of week 1.
→ Week 2-5: increase dose by 25mg/day.
→ Week 5: Consultant-guided increments of 25-50mg/day until a stable dose is reached, aiming for therapeutic plasma levels (0.35-0.5mg/l).

###### Monitoring includes

a. Physical health checks include pulse, postural BP, temperature, respiratory rate, oxygen saturations:
  → Week 1 and 2: 3 times per week
  → Week 3 and 4: 2 times per week
  → Week 5 and 6: 1 time per week
b. Haematological monitoring:
  → Troponin-I and CRP weekly for 6 weeks
  → FBC weekly until week 18, then fortnightly until week 52, then monthly onwards
c. Side-effects checklist at the same time as the physical health checks, including sedation, hypersalivation, constipation and any signs of immunosuppression or cardiac strain.

### Data Collection

This project was approved by the Oxford Health NHS Foundation Trust Audit Management and Tracking service with further ethical approvals not required. All clozapine initiations that had occurred within the four teams during the study period were obtained from the Denzapine Monitoring Service. For each patient initiated on clozapine a review of electronic health records was performed to ascertain location of clozapine initiation. Additional information gathered included demographics, date of initiation (derived from the central non-rechallenge database (CNRD) date), and primary diagnosis. Patients with no prior exposure to clozapine were classified as first-time titrations; those that had been titrated previously were classified as retitrations. For patients seen by TUNE-UP a range of clinical measures were also obtained at baseline and, where possible, at discharge (listed in Box 2).

### Statistical Analysis

Poisson regression (implemented using the statsmodels package 0.14.4 in Python 3.12.8)(23) was used to compare titration event counts in the period of interest (i.e. the 12 months when TUNE-UP was in operation), against the remaining time period. Events were analysed on a per annum basis, with the event count during the final 4-month period (October 2024 - January 2025) adjusted pro rata.

The primary analysis examined the impact of the TUNE-UP service on overall (i.e. first-time and retitration) community titration rates. Secondary analyses examined the impact of the service on combined titration rates (i.e. inpatient and community), inpatient titration rates, and on first time-titrations and retitrations separately. Exploratory analyses used paired t-tests to assess the change in clinical measures between baseline and discharge for those initiated on clozapine by TUNE-UP.

## Results

54 patients belonging to the 4 AMHTs of interest were recorded on the Denzapine Monitoring System as having potentially commenced clozapine treatment between September 2021 and January 2025. Three of these cases were registered but not actually titrated (due to the patient relocating to the area). Of the remaining 51 cases, 29 were identified as first-time titrations and 22 as retitrations. The 5 cases initiated in the final 4 months analysed were pro rata adjusted by a factor of 3 to create a Year 4 projection (taking overall N to 61). Demographic details for all patients receiving clozapine initiation are reported in Table 1 and clinical details for TUNE-UP patients are reported in Table 2.

**Table 1:**
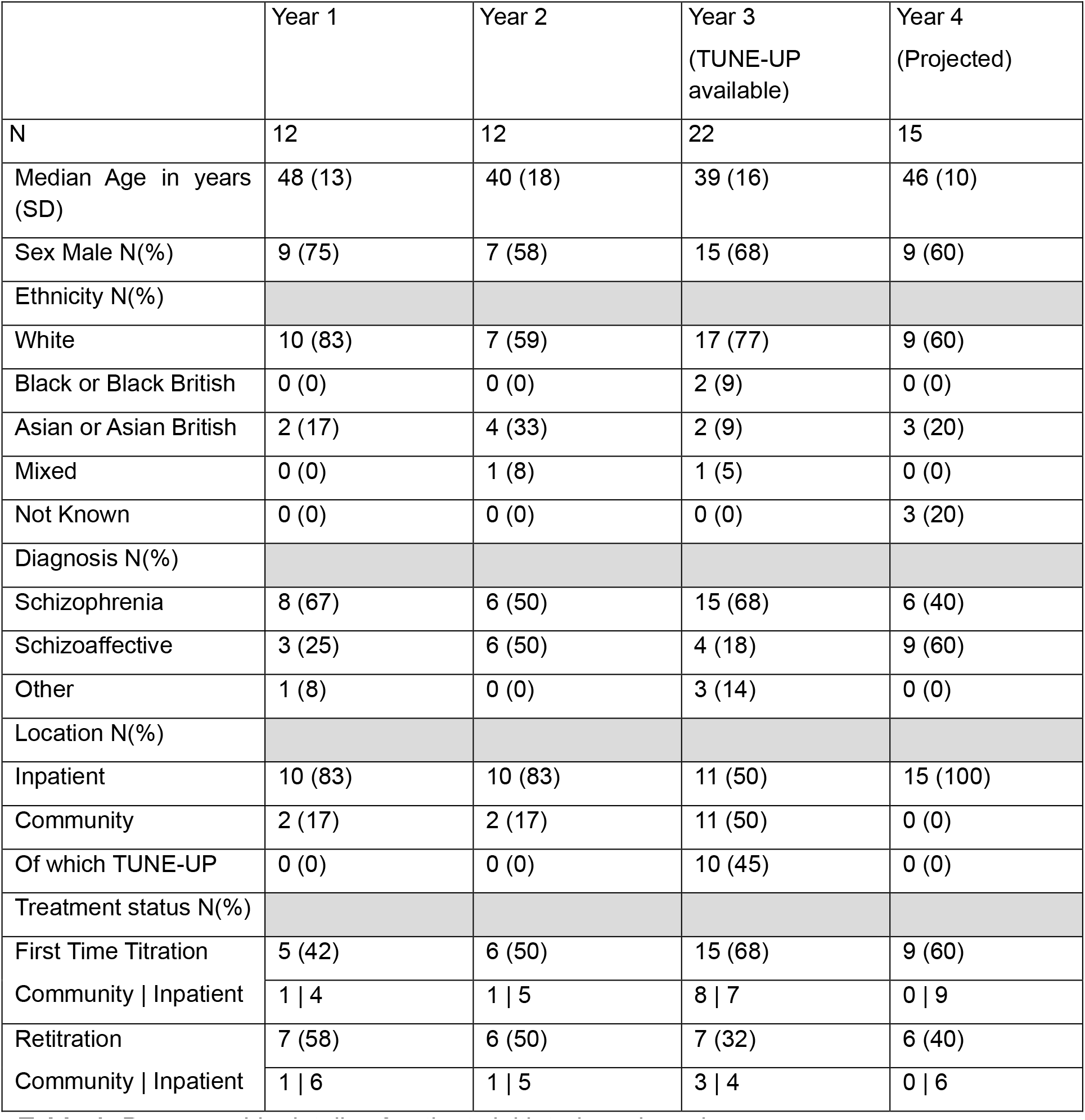
Demographic details of patients initiated on clozapine.

**Table 2:** TUNE-UP Patients Clinical Outcomes. PANSS: Positive and Negative Syndrome Scale; BNSS: Brief Negative Syndrome Scale; CDS: Calgary Depression Scale; SCI: Sleep Condition Indicator; SSTICS: Subjective Scale to Investigate Cognition in Schizophrenia; SCIP: Screen for Cognitive Impairment in Psychiatry; SOFAS: Social and Occupational Functioning Assessment Scale; SDS: Sheehan Disability Scale. ^a^Discharge scores only obtained for 6 participants. For CDS, SCI, and SDS discharge scores only obtained for two participants and so not analysed.

|  | All referrals | Clozapine<br>Initiations<br>(baseline) | Clozapine<br>Initiations<br>(change)<sup>a</sup> | Paired t-test:<br>T-statistic,<br>p-value |
| --- | --- | --- | --- | --- |
| N | 47 | 10 | 6 |  |
| Age in years (SD) | 43.6 (13.9) | 45.1 (15.8) | - |  |
| Sex Male N (%) | 35 (74) | 9 (90) | - |  |
| Referral Reason:<br>Positive<br>Symptoms N (%) | 24 (51) | 6 (60) | - |  |
| Referral Reason:<br>Negative<br>Symptoms N (%) | 1 (2) | 0 (0) | - |  |
| Referral Reason:<br>Cognitive<br>Symptoms N (%) | 3 (6) | 0 (0) | - |  |
| Referral Reason:<br>Mixed Symptoms<br>N (%) | 7 (15) | 1 (10) | - |  |
| Referral Reason:<br>Medication<br>Review N (%) | 12 (26) | 3 (30) | - |  |
| PANSS Total<br>Mean (SD) | 67.9 (20.7) | 63.3 (18.3) | -20.5 (12.2) | 4.123,<br>0.009* |
| PANSS Positive<br>Mean (SD) | 17.2 (5.8) | 25.0 (7.1) | -7.3 (6.3) | 2.859,<br>0.035* |
| BNSS Mean (SD) | 18.7 (16.7) | 21.7 (14.5) | -15.6 (12.7) | 2.738,<br>0.052 |
| CDS Mean (SD) | 4.8 (5.1) | 7.0 (8.5) | -2.5 (3.5) | - |
| SCI Mean (SD) | 19.3 (9.1) | 18.0 (7.1) | 10.5 (4.9) | - |
| SSTICS Mean<br>(SD) | 26.7 (11.6) | 30.8 (4.6) | -9.5 (6.3) | 3.256,<br>0.023* |
| SCIP Mean (SD) | 55.2 (14.2) | 62.2 (13.3) | 4.0 (5.2) | -1.879,<br>0.119 |
| SOFAS Mean<br>(SD) | 51.7 (14.5) | 48.3 (7.5) | 8.8 (7.1) | -3.055,<br>0.028* |
| SDS Mean (SD) | 18.4 (7.2) | 18.5 (10.6) | -0.5 (0.7) | - |

During the period when the TUNE-UP clozapine service was operational, clozapine initiation rates were 22.0/year (11.0/year community, 11.0/year inpatient). This compared to an average of 13.0/year (1.33/year community setting, 11.67/year inpatient), when the service was not running. This increase during the TUNEUP period was statistically significant for both the community (p = 0.0003) and total (p = 0.048) initiation rates (see Fig 1). There was no significant change in the rate of overall inpatient clozapine initiations in the time this service was running (p = 0.865).

**Figure 1:**
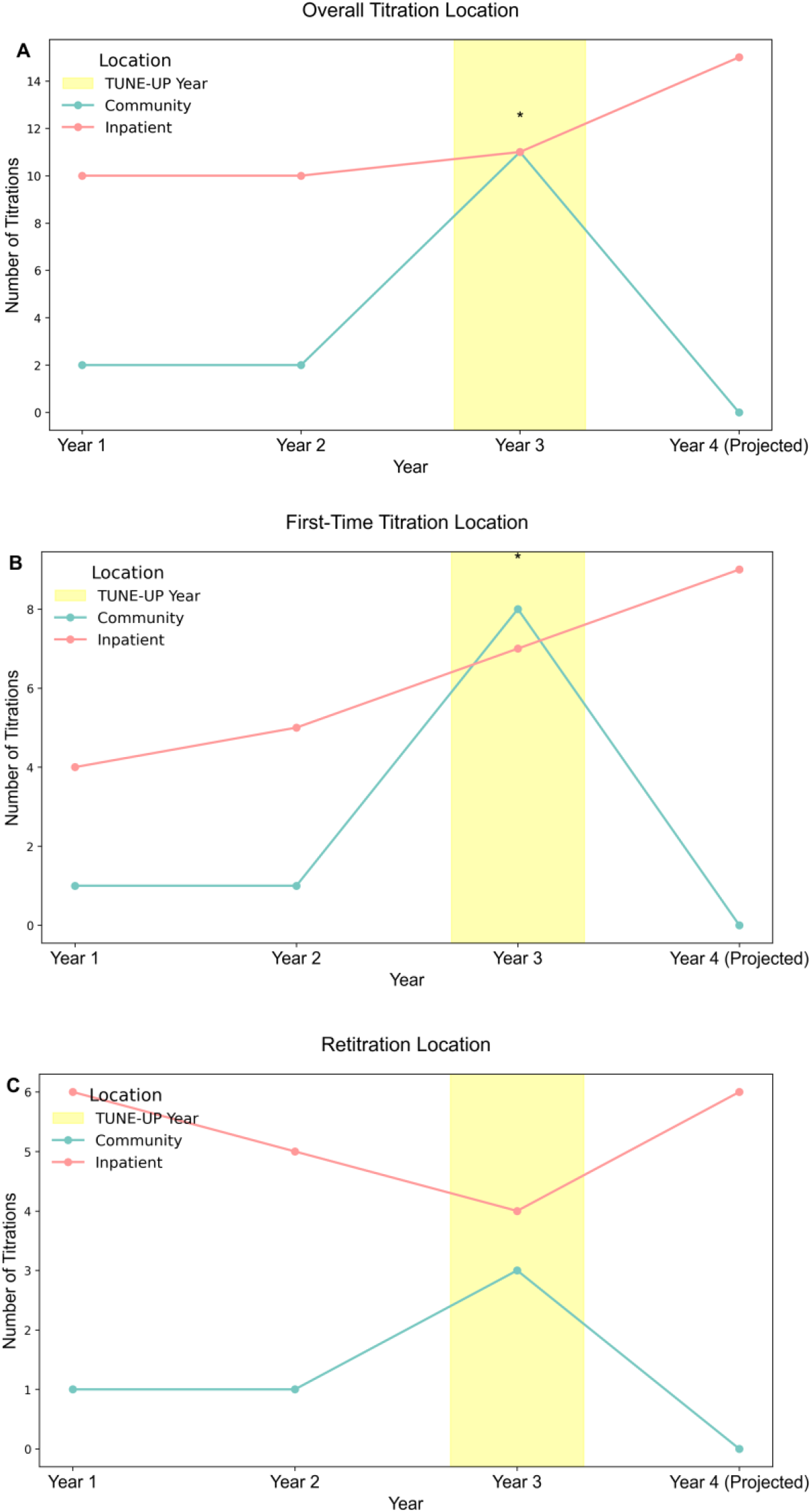
TUNEUP associated with increased community clozapine initiations In the year the TUNE-UP service was operational, there was (A) A significant increase in overall (i.e. first-time and retitration) clozapine initiations in both community (p = 0.0003) and combined (community and inpatient; p = 0.048) settings. (B) A significant increase in first-time clozapine initiations in both community (p = 0.002) and combined (community and inpatient; p = 0.018) settings. (C) No significant change in clozapine retitration rates in either community (p = 0.099) or combined settings (community and inpatient) (p = 0.821).

We next examined first-time titrations and retitrations separately. For first-time titrations there was a significant increase in the rate of community (p = 0.002) and combined (p = 0.018) titration rates, but not inpatient (p = 0.729) titrations, during the period TUNE-UP was operational. For retitrations there was no significant change in the rate of combined (p = 0.821), community (p = 0.099), or inpatient (p = 0.531) titrations during the period TUNE-UP was operational.

Of the 10 patients titrated on clozapine by TUNEUP, 6 had symptoms scored at discharge. In exploratory analyses there was a statistically significant improvement observed for total PANSS, the positive symptoms subscale, self-rated cognition, and overall functioning (see Table 2).

## Discussion

Our findings show that a targeted outpatient service for community clozapine initiation is associated with a significant increase in clozapine initiation rates. This increased rate of initiations primarily reflected patients who had not been previously titrated on clozapine (i.e. first-time titrations). There was no significant change in the rate of overall inpatient clozapine initiations in the period the service was running. While previous work has demonstrated the cost-effectiveness of community clozapine services (11), to our knowledge this is the first demonstration that specialist community initiation clinics are associated with an increase in the frequency of clozapine initiation.

Clinical guidelines advise offering clozapine to TRS patients at the earliest opportunity, and a longer delay negatively impacts prognosis.(3,4,24,25) The cost-effectiveness of clozapine treatment in TRS has been repeatedly shown, as have benefits on life expectancy and suicidality.(24,26–28) Despite this, clozapine is underutilised, and delays in initiation average five years in the UK.(29,30) There are significant differences in the rates of clozapine prescribing in different trusts, with a study in 2007 showing a persistent 5-fold difference in rates of clozapine prescription between 45 NHS trusts.(31) A more recent study in 2021 indicated that clozapine is grossly underutilised in the UK, with only one-third of eligible patients receiving this treatment, and a three-fold variation in clozapine prescription in England.(32) A previous study has shown a point prevalence of 56% of all patients under a general community mental health service meeting criteria for TRS, indicating a high rate of resistance in the community setting; half of these patients had never been trialled on clozapine.(33) This reflects similar findings in Europe and internationally.(34,35)

The underutilisation of clozapine results from a wide range of factors. Clinicians struggle to identify patients with TRS, and limitations of resources and clinicians’ and patients’ attitudes towards clozapine also play a significant role.(12,36). Practitioners’ anxiety regarding medical complications related to clozapine, concerns regarding concordance and comorbid medical conditions, the logistics of initiation, and patients’ concerns regarding tolerability and difficulty in adhering to blood test monitoring, are all barriers to timely treatment. Dedicated outpatient services focused on clozapine initiation address these barriers and are identified by practitioners as a factor that would increase prescribing,(36) and patients describe the necessity for hospital admission as the greatest barrier to agreeing to treatment with clozapine.(12) The average cost per bed day in an acute inpatient adult mental health ward of the study NHS trust is £729, and inpatient admissions for clozapine titrations last on average 100 days. The costs of the 10 additional titrations performed by TUNEUP, therefore, might be expected to have exceeded £700,000 if undertaken in an inpatient setting. This highlights how innovative community approaches are required if clozapine initiation rates are to be increased in a cost-effective manner.

The impact of the service was primarily in increasing community rates, rather than reducing inpatient admission rates. Many inpatient initiations are of patients that are significantly unwell to the extent that necessitates admission, and as such may not be suitable for community initiation. It appears that when the service was unavailable, patients were primarily offered clozapine initiation during periods of acute symptomatic exacerbation requiring inpatient admission. Therefore, the group most likely to benefit from services such as TUNE-UP may be those with severe symptoms that impair functioning, but do not require inpatient admission. With restrictions of resources contributing to a lack of inpatient beds, and limited capacity of outpatient teams to initiate clozapine, it is likely that this group would continue to be sub-optimally treated in the community, and either continue in this way or deteriorate significantly enough to warrant inpatient admission. The relative proportion of first-time titrations was at its highest in the year this service was running, indicating clinicians were able to trial clozapine in a greater number of TRS patients. An outpatient clozapine titration service may give clinicians the ability to offer clozapine to TRS patients who would not otherwise receive this treatment unless they deteriorated to the point of needing inpatient admission.

A high proportion of patients discontinue treatment with clozapine, and one of the factors previously shown to contribute to this is patients’ feelings of lack of agency regarding decisions related to their treatment and their health (37,38). Shared decision-making and person-centred care lead to improved adherence and outcomes.(38–41) Community titration may allow patients greater autonomy, which may have positive effects on overall outcomes.

Given the persistent underutilisation of clozapine, national efforts to increase patient and clinician awareness may be indicated to increase the appropriate prescription of this medication. Clearer pharmacological records of previous antipsychotics trialled, including dose, duration and monitoring of adherence (e.g. using measurement of antipsychotic levels) will help clinicians identify TRS as soon as criteria are fulfilled.(42) Replicating the findings of this study in other trusts will also contribute to their robustness.

### Strengths and Limitations

Methodological strengths of this study include the ability to identify a comprehensive cohort of individuals commencing clozapine for the first time during the study period, and the ability to accurately identify their titration location from electronic records with minimal confounders of this data. The risk of selection bias of patients in this retrospective study was limited by using a centralised registry to identify all cases of clozapine initiation. In addition, the fact that the service was piloted for 12 months, and data was gathered following the termination of the pilot provides a natural experiment which demonstrates the findings are not reflective of more general long-term trends unrelated to the service.

A limitation of this study was the moderate total number of patients, and the observational nature is unable to ascribe causality to the effect of the intervention. A further possible limitation of this study was data incompleteness: in the case of retitrations it is possible that in cases where only a short break had occurred, the retitration may not have been recorded on the DMS database, and they would therefore have been missed in the analysis. This would not however affect the analysis of first-time titrations which demonstrated consistent results with the overall findings. The robustness of the clinical change measures is limited by the fact they were obtained by unblinded clinicians and only in a small number of patients.

## Conclusion

The findings of this study demonstrate that a targeted service that can titrate patients on clozapine in the community has a significant impact on the clozapine initiation rates. There was a significant increase in both the combined rates of clozapine initiation, and the community rate when analysed separately, but not the rate of inpatient titrations. There was also a significant increase in the rate of combined and community first-time titrations when analysed separately. Patients titrated on clozapine through the service had improved symptom burden and functioning. There is now clear evidence that services for the community initiation of clozapine, which are potentially more cost-effective, increase rates of clozapine initiation and provide clinical benefit to patient groups that are otherwise being treated sub-optimally in the community.

## Acknowledgements

RAMs work is funded by a Wellcome Trust Clinical Research Career Development Fellowship (224625/Z/21/Z). This work and the TUNE UP clinic have been developed as part of a collaborative working partnership between Oxford Health NHS Trust and Boehringer Ingelheim Ltd. VM is supported by the Swiss National Science Foundation (P500PM_217669). TP is supported by the UK National Institute for Health Research (NIHR), Maudsley Charity, the Brain and Behaviour Research Foundation, the UK Academy of Medical Sciences, and the UKRI Hub for Metabolic Psychiatry (grant reference MR/Z503563/1, platform grant code MR/Z000548/1). JBF is supported by an NIHR Academic Clinical Fellowship (ACF-2023-13-016).

## Transparency Statement

RAM has received speaker/consultancy fees from Boehringer Ingelheim, Janssen, Karuna, Lundbeck, Newron, Otsuka, and Viatris, and co-directs a company that designs digital resources to support treatment of mental ill health. TP has received speaker/consultancy fees from Boehringer Ingelheim, Recordati, Lundbeck, Otsuka, Janssen, CNX Therapeutics, Sunovion, ROVI Biotech, Schwabe Pharma, and Lecturing Minds Stockholm AB; he receives book royalties from Wiley Blackwell; he co-directs a company that designs digital resources to support treatment of mental illness. Other authors have no conflicts of interest to declare.

## Author Contribution

ZAK and IV wrote the initial draft of the manuscript with RAM’s supervision. All collaborators reviewed the manuscript and provided input on the final draft.

## Data Availability

Data available from authors on request

## Notes

### Author Declarations

This project was approved by the Oxford Health NHS Foundation Trust Audit Management and Tracking service with further ethical approvals not required.

